# Proteome analysis of platelets in Essential Thrombocythemia couples metabolism to platelet reactivity

**DOI:** 10.1101/2025.03.10.25323560

**Authors:** Xiomara Guerrero-Carreño, Sanne Smits, Alfonso Esteban Lasso, Martina Samiotaki, Virtu Calabuig Navarro, Francisco José Iborra, Frida Rantanen, Alberto Alvarez-Larrán, Anna Angona Figueras, Beatriz Bellosillo, Adolfo J. Sáen Marín, Valentin García Gutiérrez, Dick Dekkers, Jeroen Demmers, Francisca Ferrer-Marín, Juan Carlos Hernández-Boluda, Antonios Matsakas, M Celina, Benavente Cuesta, Peter Vandenberghe, Petros Papadopoulos

**Affiliations:** Department of Hematology, Cellular differentiation and gene expression, Instituto de Investigación Sanitaria San Carlos (IdISSC), Hospital Clínico San Carlos, Madrid, Spain; Department of Hematology, Hospital Clínico San Carlos, Madrid, Spain; Center for Human Genetics, KU Leuven and University Hospitals Leuven, Leuven, Belgium; University Rey Juan Carlos, Madrid, Spain; Institute for Bioinnovation, BSRC “Al. Fleming”, Vari, Greece; Biological Noise and Cell Plasticity, IBV (CSIC), Valencia, Spain; Medical Systems Biology, University of Helsinki, Helsinki, Finland; Department of Hematology, Hospital del Mar, Barcelona, Spain; Department of Hematology, Clinic Hospital, Barcelona, Spain; Instiut Català d’Oncologia - Hospital Trueta Girona; Instituto Ramón y Cajal de Investigación Sanitaria, Universidad de Alcala, Spain; Proteomics Center, ErasmusMC, Rotterdam, The Netherlands; Department of Hematology, Hospital General Universitario Morales Meseguer, CIBERER, IMIB, UCAM, Murcia, Spain; Department of Hematology, Hospital Clínico Universitario, INCLIVA, Valencia, Spain; Centre for Biomedicine, Hull York Medical School, UK; Department of Hematology, University Hospitals Leuven, Leuven, Belgium

**Author notes:** **Correspondence:** Prof. dr. Peter Vandenberghe, UZ Leuven | campus Gasthuisberg | Herestraat 49 | 3000 Leuven – Belgium, Dr. Petros Papadopoulos, Medical Systems Biology, Faculty of Medicine, University of Helsinki, Finland, Haartmaninkatu 8, 00290 Helsinki. These authors contributed equally to this work (Peter Vandenberghe, Petros Papadopoulos). **Competing interests:** The authors declare to have no competing financial interests.

## Abstract

Platelets are key players in hemostasis and thrombosis. Essential thrombocythemia (ET) is a myeloproliferative neoplasm (MPN) in which the *JAK2* V617F, *MPL* W515K/L, and *CALR* mutations determine differences in clinical phenotype, in particular the thrombotic risk and the risk of myelofibrosis. Here, we examined the proteome of platelets in ET by mass spectrometry (MS) in combination with functional assays to gain insights into platelet activation in ET.

MS analysis revealed a different proteome in ET platelets with stoichiometric differences in mitochondrial proteins compared with normal platelets. The tricarboxylic acid cycle enzymes (TCA) were in general downregulated in ET platelets while glycolysis enzymes were upregulated changing modes in energy production. Acetyl salicylic acid (ASA) treatment increased levels of TCA enzymes in controls and restored them only partially in *JAK2* V617F platelets. Interestingly, membrane CD36 was higher in *CALR* Type1 implicating lipid transport and fatty acid oxidation in platelet lifespan. Aggregation levels specifically in *JAK2* V617F platelets were similar or lower to healthy controls while activation markers i.e. CD62P were higher in untreated *CALR* Type2 than controls and the rest of ET.

In summary, analysis of platelet proteome in ET implicates mitochondrial activity in platelet activation and also identified differences between *JAK2* V617F and *CALR* patients. Our study suggests that metabolic finetuning can be critical in the control of platelet reactivity.

**Key points:**

1. TCA cycle enzymes are downregulated in ET platelets. ASA treatment leads to a partial correction in *JAK2* V617F platelets.
2. CD62P expression and aggregation levels of untreated CALR Type2 are higher than *JAK2* V617F platelets.

**Visual Abstract:** 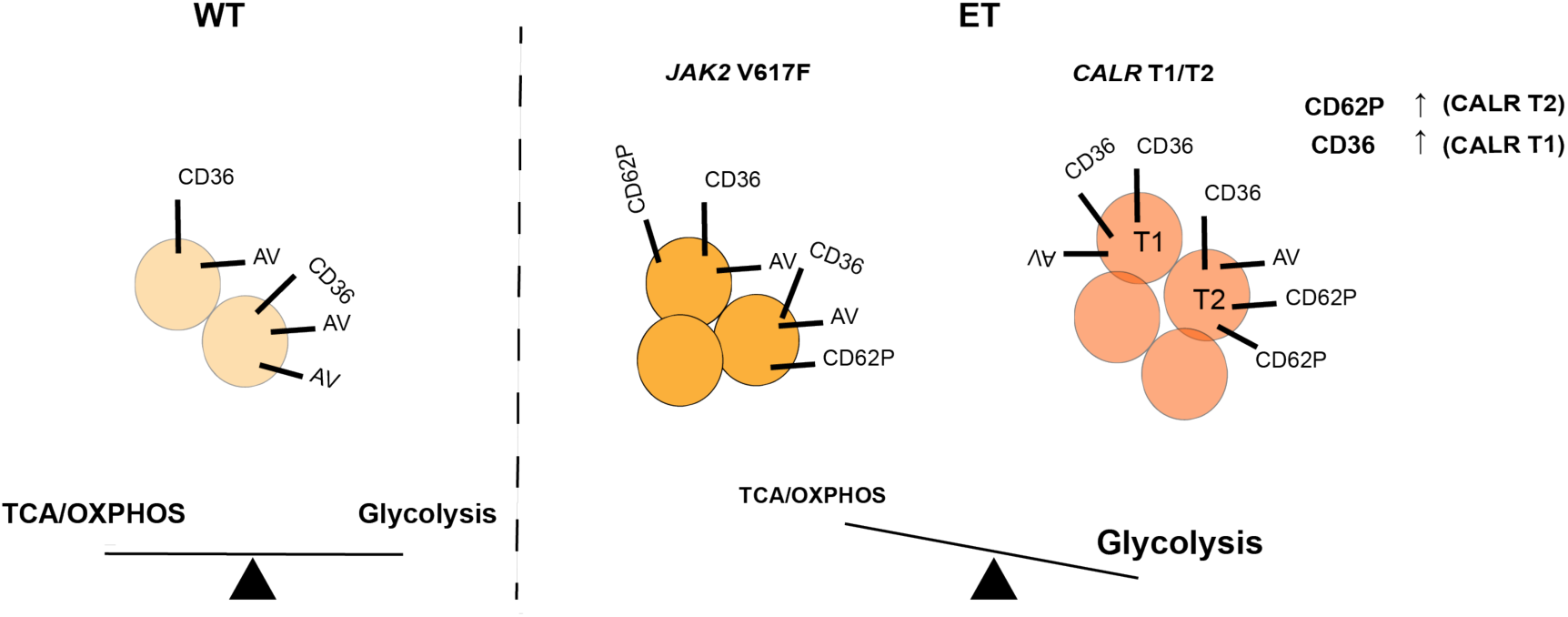

## Introduction

Patients suffering from ET are at variable risk of hemorrhagic or thrombotic complications. Their treatment is based on risk assessment and will be expectative, anti-platelet agents alone (i.e. ASA), or in combination with cytoreductive agents such as hydroxycarbamide (HU) or anagrelide (ANG) (1).

*JAK2* V617F positive ET has a higher risk of thrombotic events than ET with CALR mutations (2), which instead carries a higher risk of evolution to myelofibrosis (3). At the same time, the bleeding tendency of ET patients might be higher than previously thought, and could be increased by antiplatelet agents in both high risk (4) and low-risk patients (5).

Platelets are key players in the tissue homeostasis, and an elevated platelet count is the cardinal finding in ET. Still, our understanding of platelet activation in ET remain incomplete, especially its modulation by driver mutations and current treatment regimens. Human platelets retain their protein synthesis capacity and have a diverse transcriptome. They also retain a functional spliceosome that can process pre-mRNAs to mature transcripts so that the platelet transcriptome can change dynamically without new transcription. Additionally, platelet’s translation is controlled by the mTOR pathway(6) that is a main regulator of cell cycle progression and cell growth (7), linking the activation of platelets with the demand for energy production and metabolism. Interestingly, previous reports have linked metabolism to the harnessing of thrombosis (8, 9).

In this study we performed mass spectrometry (MS) and flow cytometry based functional assays on platelets from patients with ET and different driver mutations. We found differences between ET and normal controls, and between the *JAK2* V617F and *CALR* mutational groups, in the abundance of mitochondrial proteins, platelet activation, and platelet aggregation. Our results suggest a functional link between mitochondrial activity and metabolism with platelet activation that should be addressed in future studies.

## Methods

### Human samples

Peripheral blood was collected from patients and healthy donors after informed consent according to the guidelines of the Declaration of Helsinki and the Ethical Committee of the University Hospital Leuven (study number S53745), and Hospital Clínico San Carlos (Madrid, C.P. - C.I. 16/257-E_BS). The diagnosis of ET was made according to the WHO 2016 criteria (3, 10). A total of 76 ET patient and 36 healthy controls were included. Most of the patients included in this study were treated with low dose (80-100mg) ASA once-daily: therefore, low dose ASA was also given for 3 days to healthy controls prior to blood sampling. Samples were processed within 24h after venipuncture and kept at room temperature (RT) in rotation. Pellets of platelets were snap frozen in liquid nitrogen and stored at -80 °C for mass spectrometry (MS).

### Mass Spectrometry

Liquid chromatography MS (LC-MS/MS) on platelets was performed and analyzed as described (11) (12). Results were processed statistically and visualized with the Perseus software (1.6.15.0) ^REF^(https://maxquant.net/perseus/). The raw files and processed proteomics data of two independent experiments have been deposited to the ProteomeXchange Consortium via the PRIDE (13) partner repository with the dataset identifiers PXD052171 and PXD050550.

### Western blotting

Platelets were lysed in RIPA buffer (Sigma Aldrich) supplemented with protease inhibitors (complete, Roche) and run on gradient 4-20% mini-Protean TGX Precast gels (Biorad). Proteins were transferred to PVDF membranes. After blocking with 3% milk/0,05%TBS for 1h at RT, the membranes were incubated with a primary antibody in 1%milk/0,05%TBST for 1,5 h at room temperature or overnight at 4°C, followed by a 1h incubation at RT with a secondary antibody (HRP or dye Light 600/680 conjugated). Membranes were developed with ECL (Biorad) after three washes of 10 minutes each at RT with 0,05%TBST. The antibody list is available in the supplementary data.

### Platelet phenotyping and functional analysis

Blood was collected in EDTA tubes (BD bio). Complete blood counts (CBC) were measured on Beckman Coulter hemato-counters. Platelet rich plasma separation and flow cytometry based platelet aggregation (FCA) and activation assay were performed as previously described (14). In addition, TRAP-6 (Thrombin receptor activator, BACHEM) was used at 100μM final concentration. For the staining of platelet surface markers, 10^6^ platelets were incubated with antibodies in PBS /1% BSA at RT for 15min except for CD62P, which was prepared in HEPES (+ CaCl_2_ 2mM final) and fixed in 1% formaldehyde prior to acquisition. The list of antibodies used is available in the supplementary section.

Flow cytometry was performed on LSR Fortessa-HTS or FACSCanto-II (BD Biosciences) cytometers, and data were analyzed using FlowJo™ v10.8 Software (BD Life Sciences).

### Statistical Analysis

Visualization of the flow cytometry data and statistics were performed with the R-package and GraphPad (two-way ANOVA).

## Results

### Sample size of the study

76 ET patients were recruited, 20 with the *JAK2* V617F mutation, 8 with an *MPL* W515K/L mutation, 17 with a *CALR* Type1 mutation, 9 with a *CALR* Type2 mutation, and 22 triple negative (TN) ET i.e. without any of these driver mutations. As controls, 36 healthy donors were recruited (**Table 1& Figure 1A**). Hematological parameters are shown in **Supplementary Figure1.**

**Figure 1.**
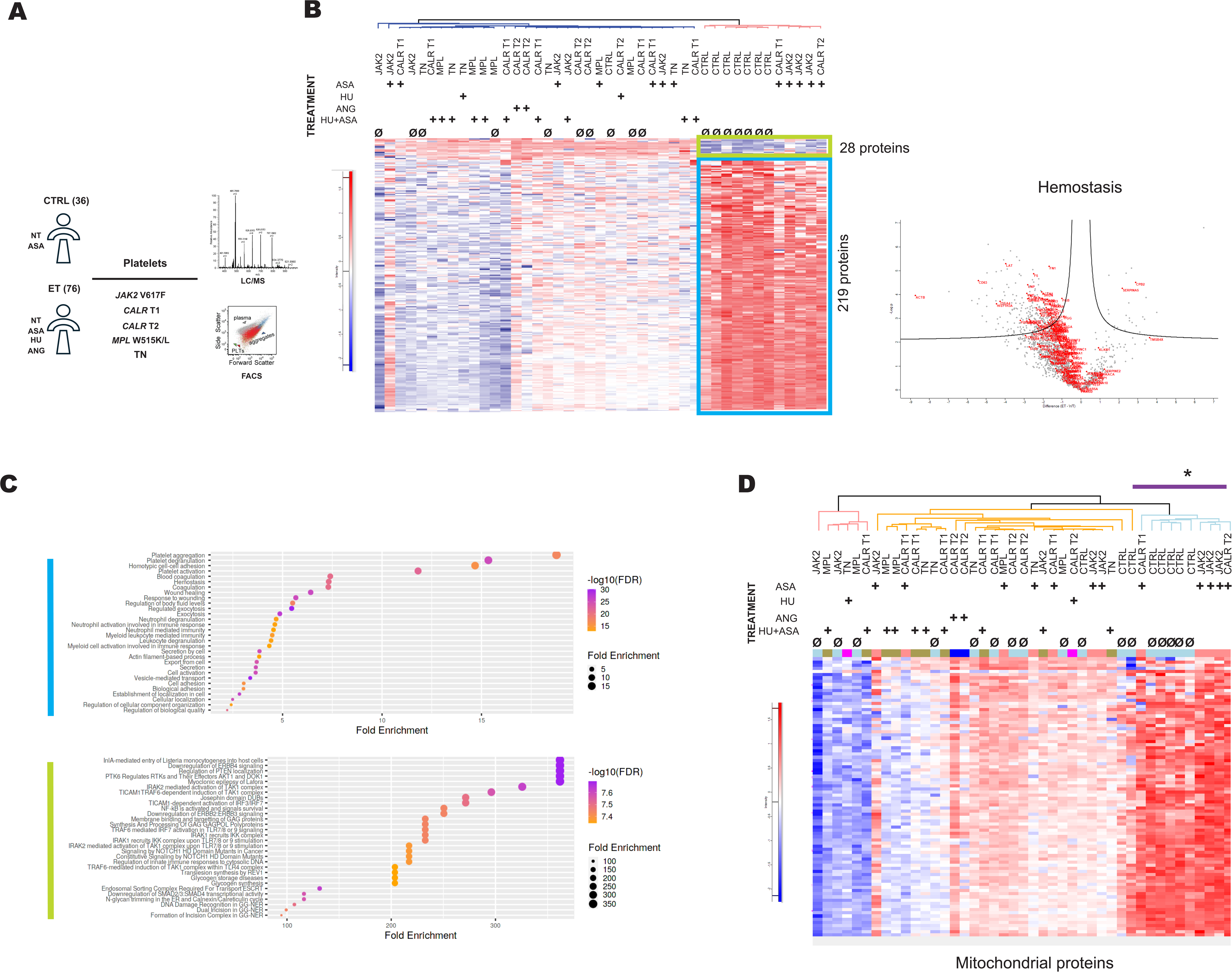
**A.** Overview of the study (NT: No treatment, ASA: acetylsalicylic acid, HU: hydroxyurea, ANG: anagrelide) **Β**. Unsupervised clustering of ET and normal control (CTRL) platelet samples after LC-MS/MS and Volcano plot of hemostasis related proteins enriched in CTRL (FDR 0.05) **C**. Pathway enrichment analysis of proteins with relative increased abundance (blue box) or relative decreased abundance (light green box) in CTRL samples shown in the heatmap (B), **D.** Unsupervised clustering of the same samples as in (A) filtered for mitochondrial proteins only. Analysis has been performed with Perseus software.

### Mass spectrometry on ET platelets

LC-MS/MS was performed on platelets from patients with ET and healthy donors, in order to get insights into their function and activation status. 43 samples were processed: *JAK2* V617F: n=9, *CALR* Type1: n=8; *CALR* Type2: n=6, MPL W515K/L: n=6, TN: n=6, and controls: n=8 (**Figure 1A**). 60 out of the 76 ET and 12 out of the 36 controls were treated with ASA, HU, ANG or a combination of them (**Table 1& Supplementary Figure 1**). The rest of them were untreated or ASA-free for controls respectively and are named as such throughout the study.

In repeated MS experiments we were unable to detect any peptides corresponding specifically to the mutant CALR using trypsin as digestion enzyme (**Supplementary Figure 2)**. We assume that mutant CALR is at very low level in platelets, as the mutant CALR (del52bp or Ins5bp) is known to be very unstable in over-expressing experiments and is rapidly degraded (15).

Unsupervised hierarchical clustering of the MS data segregated the protein signatures of ET platelets from that of healthy donors (**Figure 1B**). Healthy controls clustered together (right side of the heatmap) next to ASA-treated *JAK2* V617F samples. 219 proteins were significantly less abundant in ET platelets (FDR <0.05): with functions in hemostasis, platelet activation, aggregation and degranulation (e.g. CD63, GP1BA, GP1BB, VWF, CLEC1B), in cellular stress response (e.g. HSPA1A/B, COX3/3, BNIP2, PACS-2), in signaling by Rho GTPases and metabolic processes (e.g. ENO2, IDH2, IDH3A, COX2/3) based on GO term/KEGG and reactome gene sets (**Figure 1C**). 28 proteins (light green box) were more abundant in ET, and involved in MAP kinase activation, TGF receptor signaling, and down-regulation of EGFR/ERBB signaling (RPS27A, UBA52, UBB, UBC) (**Figure 1C**). Comparison of platelets from untreated ET and ASA-free controls showed a set of proteins related to platelet activation and aggregation (100 proteins) in lower abundance in ET indicating that the proteome differences between controls and ET samples were independent of the treatment (**Supplementary Figure 3**).

Among the 219 proteins with lower abundance in ET (**Figure 1B**) many are involved in metabolic processes and are mitochondrial. A similar clustering pattern emerges when analysis is restricted to mitochondrial proteins only (**Figure 1D**). Specifically, controls show higher abundance for approximately 88 proteins including ACO2, CS, PKM, IDH1, IDH2 etc as compared to the rest of ET samples, with the exception of *JAK2* V617F ASA-treated ET (**Figure1D**, asterisk).

In summary, the platelet proteome in ET is distinct from healthy controls with enrichment of proteins related to kinase activation indicative of an underlying inflammatory signature described previously in MPN (16, 17). The protein signatures of ASA-treated *JAK2* V617F ET were closer to healthy controls and related to platelet activation and mitochondrial function.

### Analysis of ASA-treated ET patients

ASA was the most common treatment type among the analyzed samples, and specifically in *JAK2* V617F (**Table 1**). ASA blocks the cyclooxygenase 1 (COX1) and thromboxane (TXA_2_) synthesis (18). Therefore, we analysed the proteome of the ASA-treated ET and healthy control samples only.

Proteome comparison of the ASA-treated ET versus untreated ET and ASA-free control samples unveiled a number of proteins significantly differentiating the two conditions in both *JAK2* V617F and *CALR* Type1&Type2 ET (**Supplementary Figure 4 A,B**). *JAK2* V617F samples had more proteins significantly increased in the ASA-treated condition. 2 non-treated (NT) *JAK2* V617F samples were compared to 6 ASA-treated *JAK2* V617F platelet samples (**Supplementary Figure 4A,B**) and 763 proteins (out of a total of 783) had higher abundance in the ASA-treated condition. Among them there are proteins involved in increased signaling activity (i.e. PPAR, MAPK and JAK-STAT) and many mitochondrial proteins (PDK1, ACO2, IDH2, CS, SDHA, CYC1, FH, DECR1, COX5B, GSR etc). Also, proteins related to platelet activation, aggregation and degranulation were enriched in the ASA-treated samples (CD36, CD9, GP1BA, GP1BB, PF4, GP5, GP6, GP9, VWF, SELP, MMRN1) as compared to the non-treated condition (**Supplementary Figure 4A**).

Similarly, 4 *CALR* Type1&Type2 ASA-treated samples clustered separately from the 3 non-treated *CALR*mut samples (**Supplementary Figure 4A,B**). However, the *CALR* Type1 ASA-treated platelets showed a lower abundance for 52 proteins, the majority of which are metabolic enzymes or mitochondrial proteins (e.g. TARDBP, COX5B, COX6A1, AGK), and proteins directly related to platelet activation did not significantly changed in contrast to what we observed in *JAK2* V617F ET.

Overall, the proteome analysis suggests that ASA treatment has a significant impact on metabolic processes related to mitochondria, which was stronger in *JAK2* V617F platelets and indicates a link between higher metabolic activity and platelet activation.

### Second MS experiment on platelets from ASA-treated ET and healthy controls

The lack of ASA-treated controls in the first experiment urged us to perform another MS round (LC-MS/MS) focusing on the effect of ASA and the related metabolic/mitochondrial signatures in *JAK2* V617F and *CALR* Type1 that we described above. ASA-treated *CALR* Type2 patients were rare, thus not included in this MS experiment. Three healthy controls were sampled before and after three days of low-dose ASA administration once-daily, which is sufficient time to cause COX inhibition in platelets and confer irreversible inhibition in TXA_2_ synthesis in the circulation(19). Additionally, new ET patients with *JAK2* V617F and *CALR* Type1 mutations were included. A total of 20 samples were processed for LC-MS/MS consisting of healthy controls, *JAK2* V617F, *CALR* Type1 &Type2 patients (**Figure 2A**).

**Figure 2.**
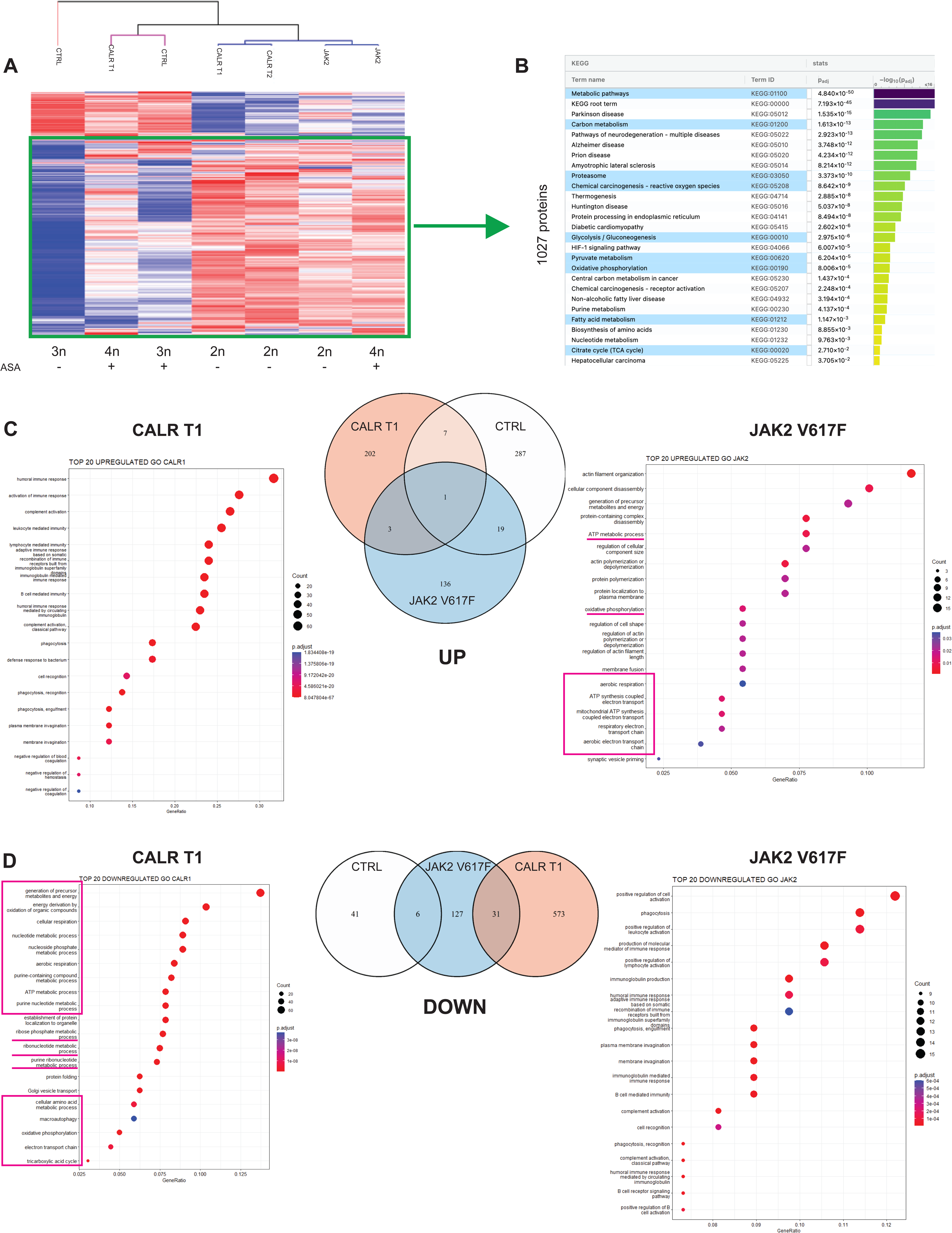
**A.** Unsupervised clustering of ET and CTRL platelet samples by MS (+/- ASA). Mean abundance values (triplicates per sample) have been generated for all proteins per mutational group (*JAK2* V617F, *CALR* Type1&Type2) and CTRL. Number of donor samples (n) per group is shown **B.** GO pathway (g:profiler) analysis on metabolic pathways from the 1027 enriched proteins (green box in **A**) **C.** Gene set enrichment analysis (GSEA) analysis of proteins with increased abundance in ASA-treatment condition for ET (*JAK2* V617F, *CALR* Type1) and CTRL samples. A Venn diagram shows the number of enriched proteins that are unique or shared on each group **D**. GSEA analysis for proteins with decreased abundance in the ASA condition for ET and CTRL platelets and Venn diagram as in **C**. (ASA: acetylsalicylic acid)

As shown in **Figure 2A** the ET samples clustered separately except from the *CALR* Type1 ASA-treated samples, which were found in between the ASA-free and ASA-treated healthy controls even though with higher abundance scores for the 1027 proteins (green box). This group of stoichiometrically enriched proteins, which were mostly shared by the ET platelets (shown in the green box of the heatmap at **Figure 2A**) were strongly related to metabolism, glycolysis, TCA and fatty acid metabolism apart from platelet activation, degranulation, aggregation and hemostasis (**Figure 2B**). Comparing ASA treatment versus ASA-free conditions we found no overlap in proteins with significant abundance changes between ET mutational groups (*JAK2* V617F, *CALR* Type1&*CALR* Type2) and healthy controls. Gene ontology (GO) analysis of the 202 proteins uniquely enriched in the *CALR* Type1 ASA-treated samples highlighted immune related processes while the respective 136 proteins in *JAK2* V617F were rather related to actin polymerization, mitochondrial ATP synthesis and oxidative phosphorylation (**Figure 2C**). Interestingly, none of these pathways were found in the 314 enriched proteins deregulated in ASA-treated healthy controls. Regarding the proteins with reduced stoichiometry in the ASA condition, 573 were found to be unique in *CALR* Type1 platelets and were related to mitochondrial and nucleotide metabolism while 127 unique proteins of the *JAK2* V617F platelets were mainly related to immune responses (**Figure 2D**).

These results emphasize the different dynamics between *JAK2* V617F and *CALR* mutated ET in the stoichiometries of mitochondrial proteins and the related metabolic pathways upon ASA treatment. *JAK2* V617F proteome was significantly enriched for several mitochondrial proteins upon ASA treatment that were not found in healthy controls and which were reduced in *CALR* Type1 platelets.

### Western blotting on platelet lysates

The differences in the abundance of mitochondrial proteins upon ASA treatment between ET platelets prompted us to investigate the metabolic pathways linked to energy production and specidically glycolysis and tricarboxylic acid cycle/oxidative phosphorylation (TCA/OXPHOS) (20) (**Figure 3A**). Protein levels for several enzymes involved in the respective pathways were assessed by western blotting in ASA-treated and ASA-free conditions.

**Figure 3.**
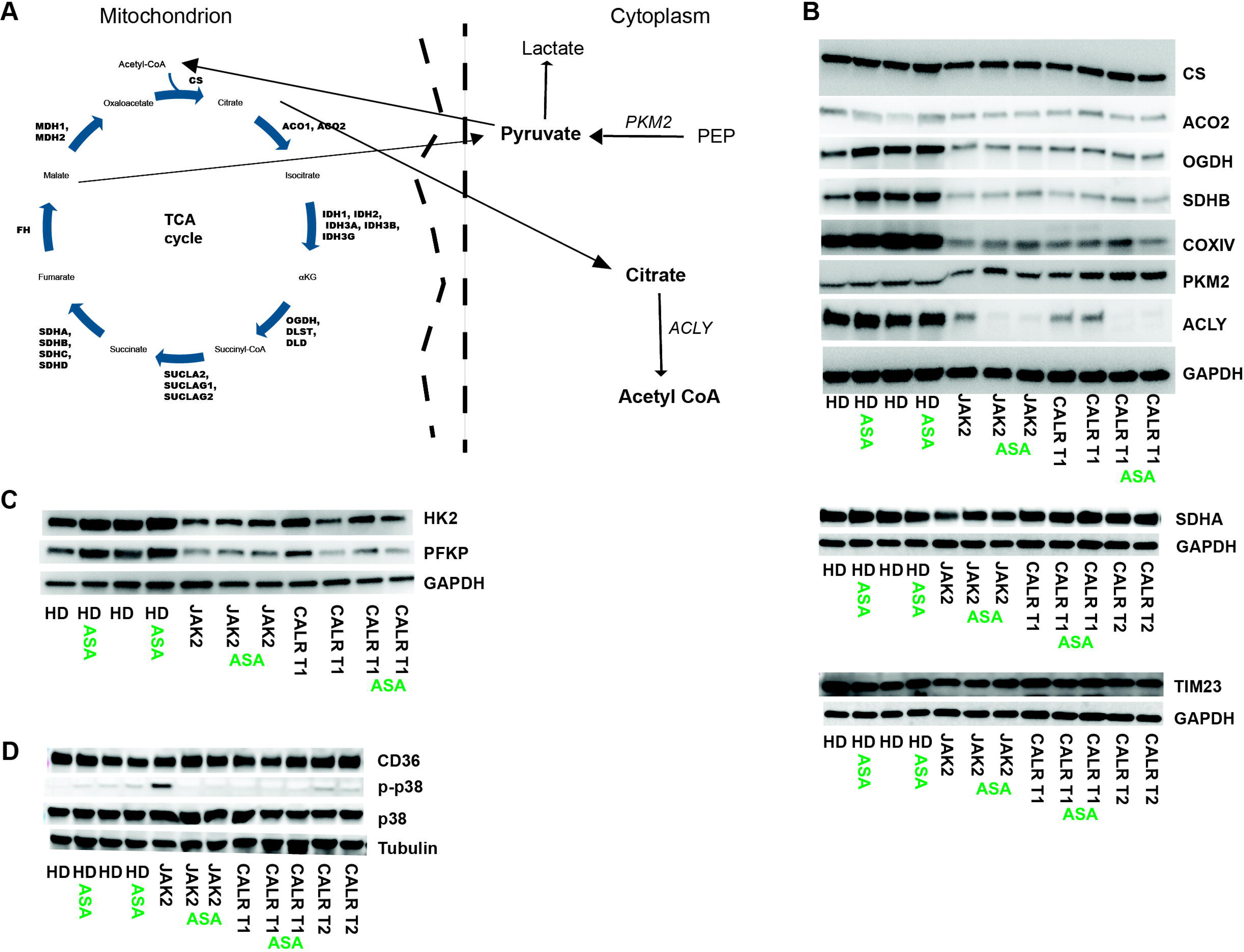
**A.** Tricarboxylic acid cycle (TCA cycle) scheme **B.** Western blot analysis on platelet lysates for the main TCA enzymes including aconitase (ACO2), citrate synthase (CS), oxoglutarate dehydrogenase complex (OGDH), succinate dehydrogenase A and B (SDHA/B), cytochrome c oxidase subunit IV (COXIV) of the electron transport chain (ETC), the pyruvate kinase PKM2 (marker for aerobic glycolysis), the cytosolic ATP-citrate lyase (ACLY) and TIM23 of the inner mitochondrial membrane **C**. Hexokinase 2 (HK2) and Phosphofructokinase, Platelet type (PFPK) are key enzymes in glycolysis **D.** Other proteins related to platelet reactivity (CD36, p38 and phospho-p38) in ET patients and CTRL (+/- ASA). GAPDH or tubulin have been used as loading controls.

All TCA enzymes and the COXIV (electron transport chain, ETC) were found at lower levels in ET platelets as compared to normal controls except from the succinate dehydrogenase A (SDHA) that was lower only in *JAK2* V617F platelets. However, levels of the inner mitochondrial membrane TIM23 (**Figure 3B**) were found to be comparable in all ET and control samples independently of treatment with ASA, which suggests that mitochondrial mass does not change significantly.

Pyruvate kinase (PKM2) catalyzes the final rate-limiting step of glycolysis by converting phosphoenolpyruvate (PEP) to pyruvate while phosphofructokinase (PFKP) and hexokinase 2 (HK2) are prominent regulatory enzymes of glycolysis. We found higher PKM2 levels in ET platelets that further increased upon ASA treatment specifically in CALR Type1 (**Figure 3B**) while PFKP and HK2 levels were reduced in ET platelets in general (**Figure 3C**). Upon ASA treatment PFKP and HK2 went up in controls and *JAK2* V617F . On the contrary, the ATP-citrate lyase (ACLY) a cytosolic enzyme that converts mitochondria derived citrate into acetyl CoA, was reduced drastically in ET (**Figure 3B**) suggesting a more glycolytic phenotype as described before in tumor cells (21). Of note, ACLY was downregulated upon ASA opposite of what was observed in healthy control platelets.

We also checked the levels of several proteins unrelated to mitochondria including p38 MAPK and CD36 described to be involved in platelet activation (22, 23) (**Figure 3D**). p38 MAPK levels remained stable, although phosphorylation of p38 MAPK was higher in ASA-free JAK2 *V617F* platelets and undetectable in ASA-treated JAK2 *V617F*. Also, CD36 was higher in *CALR* Type2 as compared to ASA-free ET and upregulated in the ASA-treated *JAK2* V617F platelets as we observed in MS (**Figure 3D).**

Taken together these results, ET platelets downregulated TCA cycle enzymes while shifting to a more glycolytic metabolism. Also, ASA treatment had a positive effect on the levels of TCA enzymes clearly seen in normal controls and to a lesser extent in *JAK2* V617F platelets in line with the proteomic analysis.

### Platelet activation

As the proteome analysis uncovered metabolic signatures that differ between *JAK2* V617F and *CALR* mutant platelets we set out to characterize their activation status in order to see whether there is any correlation with the proteome. To this end, we measured the expression of surface markers related to platelet activation including CD36 (22, 24) and CD62P, CD63 upon agonist stimulation (i.e. TRAP6, PMA, collagen) in healthy controls and ET (**Figure 4A**).

**Figure 4.**
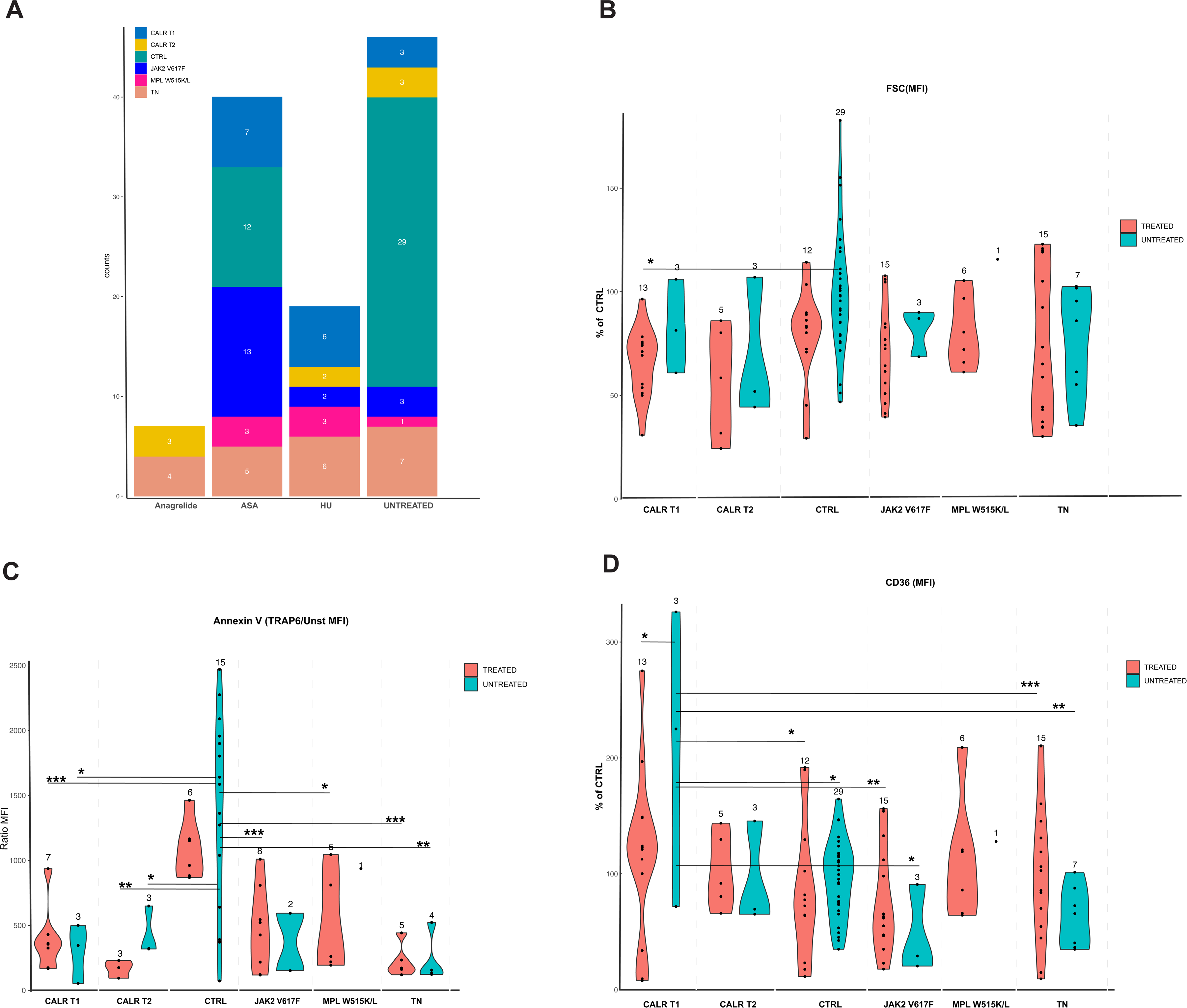
**A.** Number of samples per mutational group and treatment of ET and CTRL included in the analysis of surface marker relative expression **B.** Mean fluorescence intensity (MFI) of the Forward scatter (FSC) obtained by flow cytometry analysis. Untreated CTRL mean FSC has been set to 100 and values of all samples have been represented as a percentage of the CTRL mean MFI **C.** Ratio of MFI of Annexin-V (AV) upon TRAP6 stimulation and unstimulated condition (Unst) **D.** CD36 levels with CTRL mean MFI set to 100 as in (**B**). Treated samples (red violin graphs) include all different treatment regimens alone or in combination (ASA, HU, ANG) except from CTRL and *JAK2* V617F in which all treated samples are only ASA-treated. Non-treated samples (including ASA-free) are shown in light blue violin plots. Numbers of samples per group are indicated on top of every violin plot. Differences are shown when statistical significance was reached in equal or more than 3 samples per group analyzed (*p-val ≤0.05, **p-val ≤0.01, ***p-val ≤0.001), (CTRL: control, ASA: acetylsalicylic acid, HU: hydrocarbamide, ANG: anagrelide)

We measured forward scatter (FSC) as a relative indicator of platelet size and mean fluorescence intensity (MFI) to accurately assess the changes in expression levels of activation markers per surface area in the membrane of platelets. No major differences were found among samples in terms of size either between mutational groups or treatment types (**Figure 4B&Supplementary Figure 5A**). Mean platelet volume (MPV) did not change significantly either (**Supplementary Figure 1**). Additionally, Annexin-V (AV) levels were assessed as a marker of apoptotic cells as it recognizes phosphatidylserine (PS) exposed in the membrane upon platelet activation (25). Interestingly, we observed lower levels (MFI) of AV in ET platelets as compared to controls upon TRAP6 stimulation and specifically in the untreated condition (**Figure 4C**). A similar trend was observed upon PMA or collagen stimulation but it was not statistically significant (**Supplementary Figure 6A, B**).

CD36 levels were higher in untreated *CALR* Type1 than normal controls, *JAK2* V617F, TN and treated *CALR* Type1 platelets **(Figure 4D)**. Instead, CD62P expression levels were higher in *CALR* Type2 untreated platelets upon PMA and TRAP6 stimulation and differences were more significant in PMA (**Figure 5A,B**). Also, CD62P levels decreased significantly in treated samples only in *CALR* Type2 either with TRAP6 or PMA. With respect to CD63 levels no significant changes were observed among samples (**Supplementary Figure 6D-F**).

**Figure 5.**
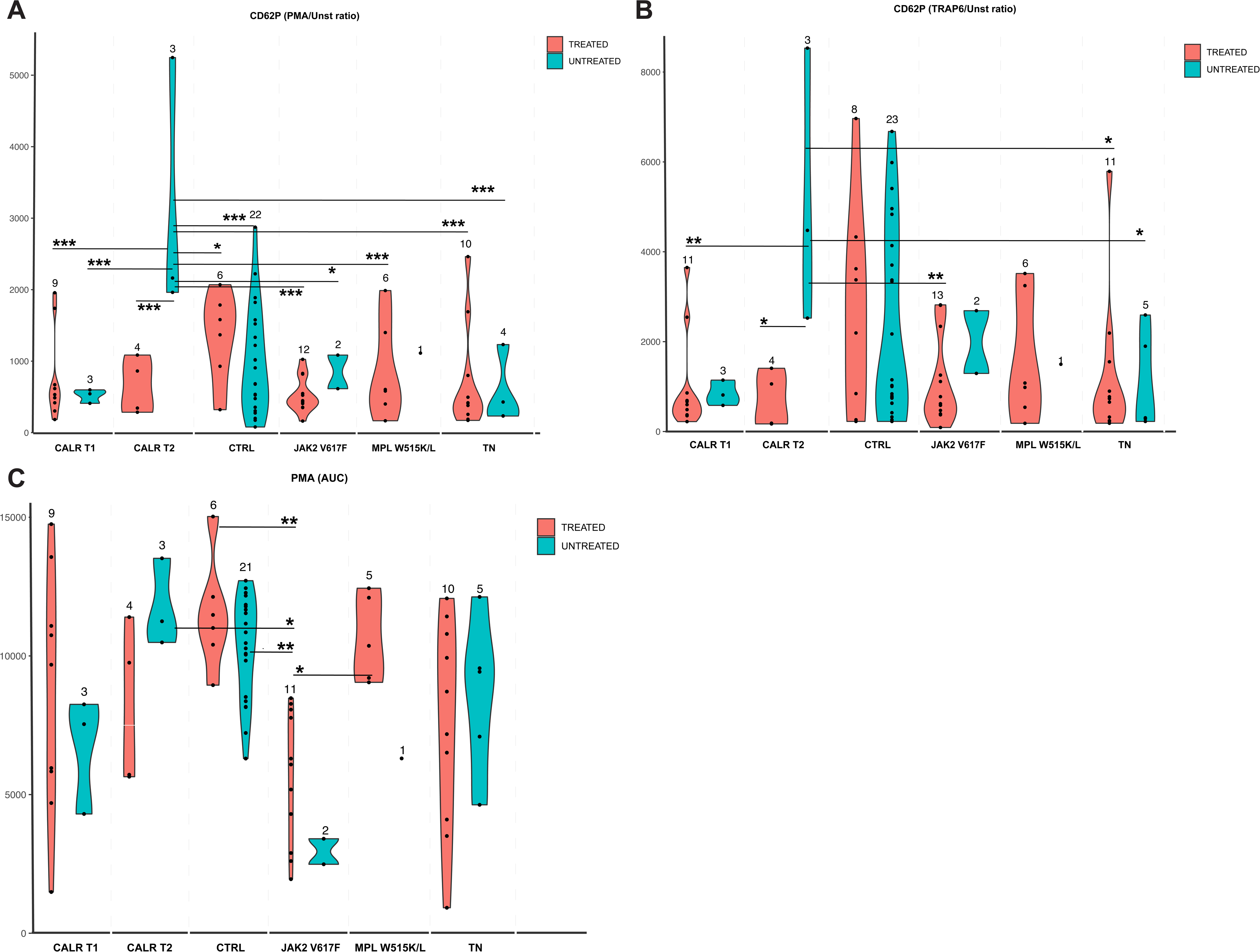
**A.** CD62P MFI ratio of relative expression levels upon PMA stimulation and unstimulated condition (Unst) and **B.** upon TRAP6 stimulation, CD62P MFI(TRAP6/Unst) **C.** Aggregation levels (area under the curve-AUC) of ET and CTRL platelets upon PMA stimulation. Treated samples (red violin plots) include all different treatment regimens alone or in combination (ASA, HU, ANG) except from CTRL and *JAK2* V617F groups in which all treated samples are only ASA-treated. Non-treated samples (including ASA-free) are shown in light blue violin plots. Numbers of samples per group are indicated on top of every violin plot. Differences are shown when statistical significance was reached in equal or more than 3 samples per group analyzed (*p-val σ0.05, **p-val σ0.01, ***p-val σ0.001), (CTRL: control, ASA: acetylsalicylic acid, HU: hydrocarbamide, ANG: anagrelide)

We also measured expression of several abundantly expressed platelet surface markers (CD49B, CD31, CD61, CD42A, CD42B, CD9, GPVI) not directly related to platelet activation. No significant differences were observed for these markers except for CD49B that was higher in controls in the ASA-treatment condition (**Supplementary Figure 5).**

Overall, ET platelets have very similar expression profiles of surface markers with the exception of CD36 and CD62P markers, which are related to platelet activation and were higher in *CALR* Type1 and Type2 respectively. Instead, *JAK2* V617F platelets presented an expression profile similar to normal controls for these membrane markers.

### Platelet aggregation

We next sought out to characterize the aggregation capacity of ET platelets by flow cytometry at a single receptor level (14). The assay is suitable for the detection and quantification even of small scale aggregates (microaggregates) and allows a dynamic analysis of the aggregation capacity of platelets during time. Six receptors were analyzed (α2α1, αII2Bβ3, GPVI, CLEC2, VWFR, and PAR1) on a time course stimulation (15sec to 10min) using specific agonists for each receptor (collagen -α2β1 and GPVI-, PMA -αIIBβ3-, convulxin -GPVI-, Aggretin A -CLEC2-, ristocetin -VWFR-, and TRAP6-PAR1).

The area under the curve (AUC) was calculated (% aggregation) on a time course agonist stimulation for each sample. Basal aggregation (% of aggregates at Time 0) and spontaneous (no agonists) aggregation (% of aggregates at Time10min) were also calculated but no significant differences were observed (**Supplementary Figure 7A,B**).

In general, ET platelets did not present significant differences in aggregation (AUC) upon stimulation with agonists as compared to healthy controls with the exception of *JAK2* V617F ET. Interestingly, *JAK2* V617F platelets showed significantly lower aggregation levels than controls (ASA condition) on PMA and TRAP6 stimulation (**Figure 5C& Supplementary Figure 7C**). No differences were found for the other agonists tested (**Supplementary Figure 7D-G**).

## Discussion

Functional studies on ET platelets have been reported in the past (26–31), many of them focusing on the *JAK2* V617F mutation. Even though the volume of the studies is growing with regard to the effects of the different treatment strategies (cytostatic or anti-platelet agents) and their potential role in the development of the most life-threatening symptoms (thrombosis or hemorrhage), there are still conceptual gaps regarding the efficacy and the metabolic rewiring upon treatment (28, 32–37). In addition, the proteome of ET platelets has not been widely explored and such analyses are very informative in order to have a comprehensive understanding of platelet biology in MPN (33, 38, 39).

In this study, 47 ET and 11 control platelet samples from treated (ASA, HU, ANG) and untreated conditions were subjected to liquid chromatography mass spectrometry (LC-MS/MS) in two separate experiments. Of note, the study of the untreated patient samples as well as ASA-treated controls emphasized the importance of treatment conditions in the control samples in MS and the functional analyses by flow cytometry. That was evident in the proteome analysis and western blotting of ASA-treated ET patients and healthy controls. *JAK2* V617F and *CALR* mutant platelets responded differently to ASA as reflected by their metabolic signatures and stoichiometries of mitochondrial proteins by MS that increased in *JAK2* V617F while mostly reduced in *CALR* Type1 (**Figure 2 & Supplementary Figure 4**).

We also checked the levels of TIM23, a stable structural mitochondrial protein and member of the presequence translocase of the inner membrane TIM23 complex (40). No significant changes were observed between control and ET platelets with or without ASA. This contrasts with a recent report (33) in which TOM20, a component of the mitochondrial outer membrane TOM complex (41), was increased in ET platelets. It has been shown previously that MPN platelets and in particular ET platelets have higher numbers of mitochondria(34) but this corresponds only to a fraction of the total numbers in the circulation due to the clonality of the disease.

Regarding the dominant bioenergetic route and its association to platelet activation in ET we have looked at the expression levels of TCA cycle and main glycolytic enzymes. The general decrease in all TCA cycle enzymes indicates that there was at least a TCA deceleration in ET platelets that was partially restored upon ASA treatment, which was detected in *JAK2* V617F platelets by the MS and verified by the western blot analysis. This is indicative of a higher metabolic flexibility, which is needed in activated platelets as it has been reported for *JAK2* V617F (26, 42). In addition, our results support previous findings that show a switch from oxidative phosphorylation to glycolysis upon platelet activation (8). Still, the source of energy production depends on the availability of metabolic blocks inside the platelets and in the circulation, which can be glucose/glycogen or fatty acids. Interestingly, the expression of CD36, a fatty acid transporter (22, 43), was higher on the surface of *CALR* Type1 platelets from untreated patients (**Figure 4D**), which suggests that there might be different levels of metabolic control that define platelet activation and/or toxicity (43) and harnessing them could prevent high platelet reactivity and thrombus formation as it has been suggested previously(9). In line with this notion similar observations have been made regarding the correction of dysregulated inflammation due to IL-10 deficiency by modulating fatty acid homeostasis (44).

Finally, *JAK2* V617F platelet activation and aggregation levels were within the range of other ET groups(45) but they were also significantly lower than healthy controls when stimulated with agonists. This was reflected in their proteomic profile, which differed from CALR mutant platelets and normal control platelets as shown upon treatment with ASA (**Figure 2**). In addition, we show that *CALR* Type1&Type2 platelet activation status is not exactly the same, as levels of CD62P were significantly higher in *CALR* Type2 untreated patients (**Figure 5A, B**).

A limitation of this study is the low number of samples per treatment regimen. However, we show that mitochondria and the associated metabolic rewiring upon platelet activation due to chronic inflammation are specific for *JAK2* V617F and *CALR* Type1. More detailed analyses on the functional status of the mitochondria are necessary to identify metabolic targets that could control platelet reactivity. This could lead to the development of more efficient therapeutic strategies in the management of MPN patients.

## Data Availability

The raw files and processed proteomics data of two independent experiments have been deposited to the ProteomeXchange Consortium via the PRIDE (13) partner repository with the dataset identifiers PXD052171 and PXD050550.

https://proteomecentral.proteomexchange.org/ui

## Acknowledgements

This study was supported by funds from the Comunidad de Madrid “Atracción de Talento” (2016-T1/BMD-1051& 2020-5A/BMD-19731 to P.P), the Stichting tegen Kanker (C/2014/302 to P.V) and Research Foundation – Flanders (FWO, G.0908.15 to P.V). We would like to thank Dr. Laura Gutiérrez (Instituto de Investigación del Principado de Asturias -ISPA-and University of Oviedo) for suggestions on the interpretation of the data and Laura García Moreno, Angela Hernández Sanchez (Hematology department of the Hospital Clínico San Carlos) and Dr. Maria Alicia Senin Magan (Hospital del Mar, Barcelona) for logistics, patient sample processing and data management. Finally, we would like to thank the healthy donors and specifically the ET patients who kindly volunteered to participate in this study.

## Authorship Contributions

X.G.C, S.S., M.S., V.C.N., D.D, performed experiments and analyzed data. A.E.L, F.R., and J.D., analyzed data. A.A.L., A.A., B.B., A.J.S.M., V.G.G., F.F.M., J.C.H.B., provided samples of ET patients, read the paper and contributed with comments. F.J.I., A.M., provided reagents, read the paper and provided comments. C.M.B.C. read the manuscript and provided comments. P.V., and P.P. designed experiments, analyzed data and wrote the paper.

