## Supplementary files for "Proteome analysis of platelets in Essential Thrombocythemia couples metabolism to platelet reactivity"

**Short title:** Proteome and functional analysis of ET platelets

Xiomara Guerrero-Carreño[1,2], Sanne Smits[3], Alfonso Esteban Lasso[4], Martina Samiotaki[5], Virtu Calabuig Navarro[6], Francisco José Iborra[6], Frida Rantanen[7], Alberto Alvarez-Larrán[8,9], Anna Angona Figueras[8,10], Beatriz Bellosillo[8], Adolfo J. Sáen Marín[11], Valentin García Gutiérrez[11], Dick Dekkers[12], Jeroen Demmers[12], Francisca Ferrer-Marín[13], Juan Carlos Hernández-Boluda[14], Antonios Matsakas[15], Celina Mª Benavente Cuesta[2], Peter Vandenberghe[3,16*], and Petros Papadopoulos[1,2,7*]

[1]Department of Hematology, Cellular differentiation and gene expression, Instituto de Investigación Sanitaria San Carlos (IdISSC), Hospital Clínico San Carlos, Madrid, Spain

[2]Department of Hematology, Hospital Clínico San Carlos, Madrid, Spain

[3]Center for Human Genetics, KU Leuven and University Hospitals Leuven, Leuven, Belgium

[4]University Rey Juan Carlos, Madrid, Spain

[5]Institute for Bioinovation, BSRC “Al. Fleming”, Vari, Greece

[6]Biological Noise and Cell Plasticity, IBV (CSIC), Valencia, Spain

[7]Medical Systems Biology, University of Helsinki, Helsinki, Finland

[8]Department of Hematology, Hospital del Mar, Barcelona, Spain

[9]Department of Hematology, Clinic Hospital, Barcelona, Spain

[10]Instiut Català d'Oncologia - Hospital Trueta Girona

[11]Instituto Ramón y Cajal de Investigación Sanitaria, Universidad de Alcala, Spain

[12]Proteomics Center, ErasmusMC, Rotterdam, The Netherlands

[13]Department of Hematology, Hospital General Universitario Morales Meseguer, CIBERER, IMIB, UCAM, Murcia, Spain

[14]Department of Hematology, Hospital Clínico Universitario, INCLIVA, Valencia, Spain

[15]Centre for Biomedicine, Hull York Medical School, UK

[16]Department of Hematology, University Hospitals Leuven, Leuven, Belgium

**Correspondence:**
Prof. dr. Peter Vandenberghe, UZ Leuven | campus Gasthuisberg | Herestraat 49 | 3000 Leuven – Belgium, tel: +32 16 34 6880

Dr. Petros Papadopoulos, Medical Systems Biology, Faculty of Medicine, University of Helsinki, Finland, Haartmaninkatu 8, 00290 Helsinki, tel: +358 29 412 5544

* These authors contributed equally to this work

### Competing interests

The authors declare to have no competing financial interests.

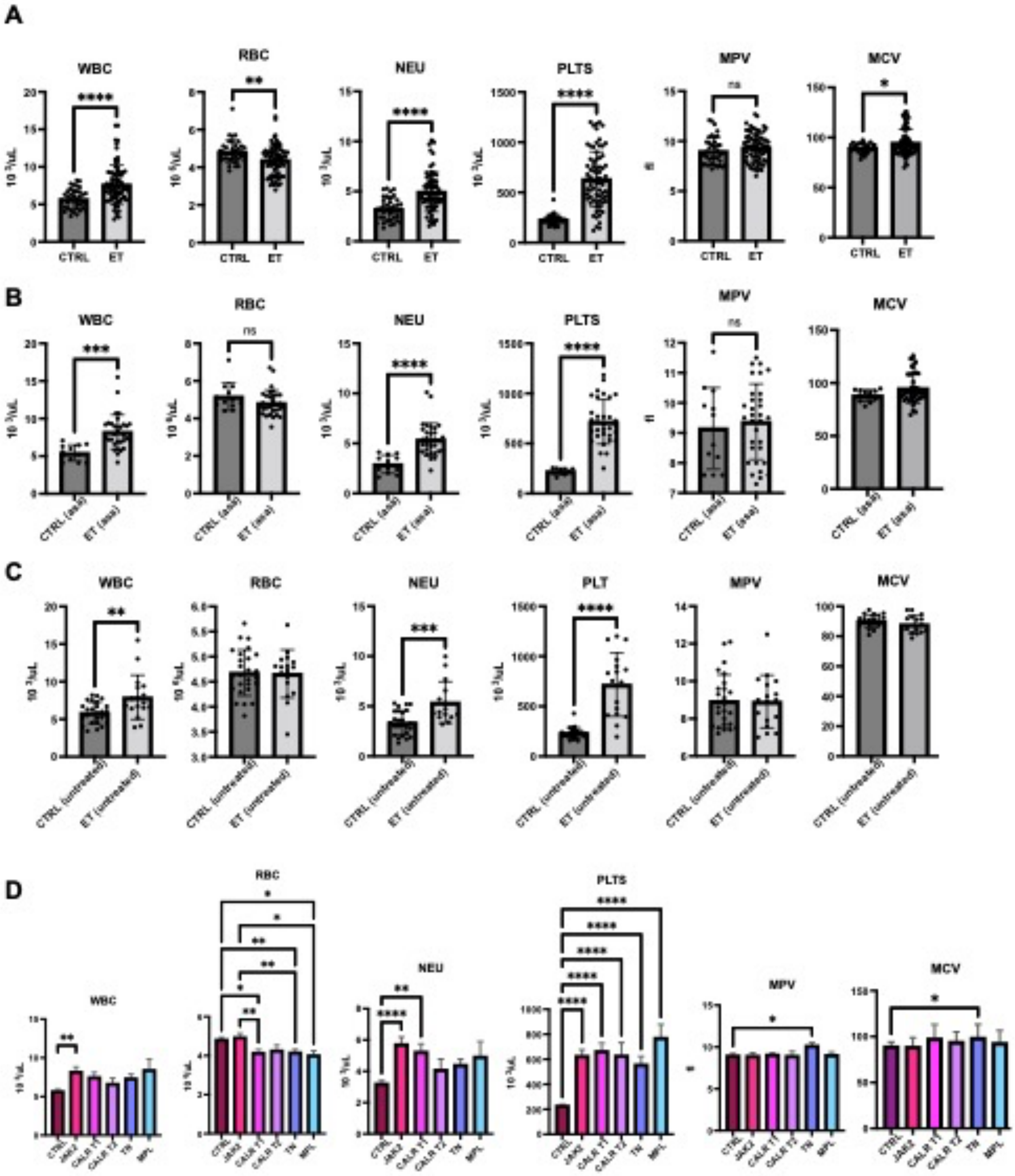


**Supplementary Figure 1**
**A.** Hematological parameters of ET patients included in the study. Data are plotted in two main groups, ET patients and healthy controls (CTRL) without any further subdivisions **B.** ASA-treated samples only **C.** Untreated samples **D**. The same parameters are shown according to driver mutation (*JAK2* V617F, *CALR* Type1&Type2, and *MPL* W515K/L) including samples with no driver mutations (triple negative, TN). WBC: White Blood Cells, RBC: Red Blood Cells, NEU: Neutrophils, PLT: platelets, MPV: Mean Platelet Volume, MCV: Mean Cell Volume.

Among the ET patients, 21% (16/76) were untreated, 42% (32/76) were receiving anti-platelet treatment (acetylsalicylic acid: ASA) and 37% (28/76) were receiving cytostatic treatment (hydroxycarbamide -HU, anagrelide-ANG), from which HU alone (21%, 6/28) and ANG alone (25%, 7/28) or in combination with anti-coagulant or anti-platelet treatment (54%,15/28). We also collected 36 control samples from healthy donors (CTRL). Since the most frequent treatment among patients was ASA (42%) we also obtained blood samples from 12 normal controls that had been on low dose ASA once-daily for 3-days prior to blood sampling **(Table 1)**.

Hematological parameters were measured on a Coulter hemato-counter. There was a significant increase in white blood cells (WBC) in untreated or ASA-treated ET patients, mainly due to increased neutrophil counts in the *JAK2* V617F and *CALR* Type1 patients (**Supplementary Figure 1A-D**). Also, there was a tendency to higher RBC counts in *JAK2* V617F ET patients as compared to the other mutational ET groups and independently of treatment (**Supplementary 1D**). The mean platelet volume (MPV) did not change significantly among ET samples except for TN, which presented higher mean cell volume (MCV) and MPV values (**Supplementary 1A, D**).

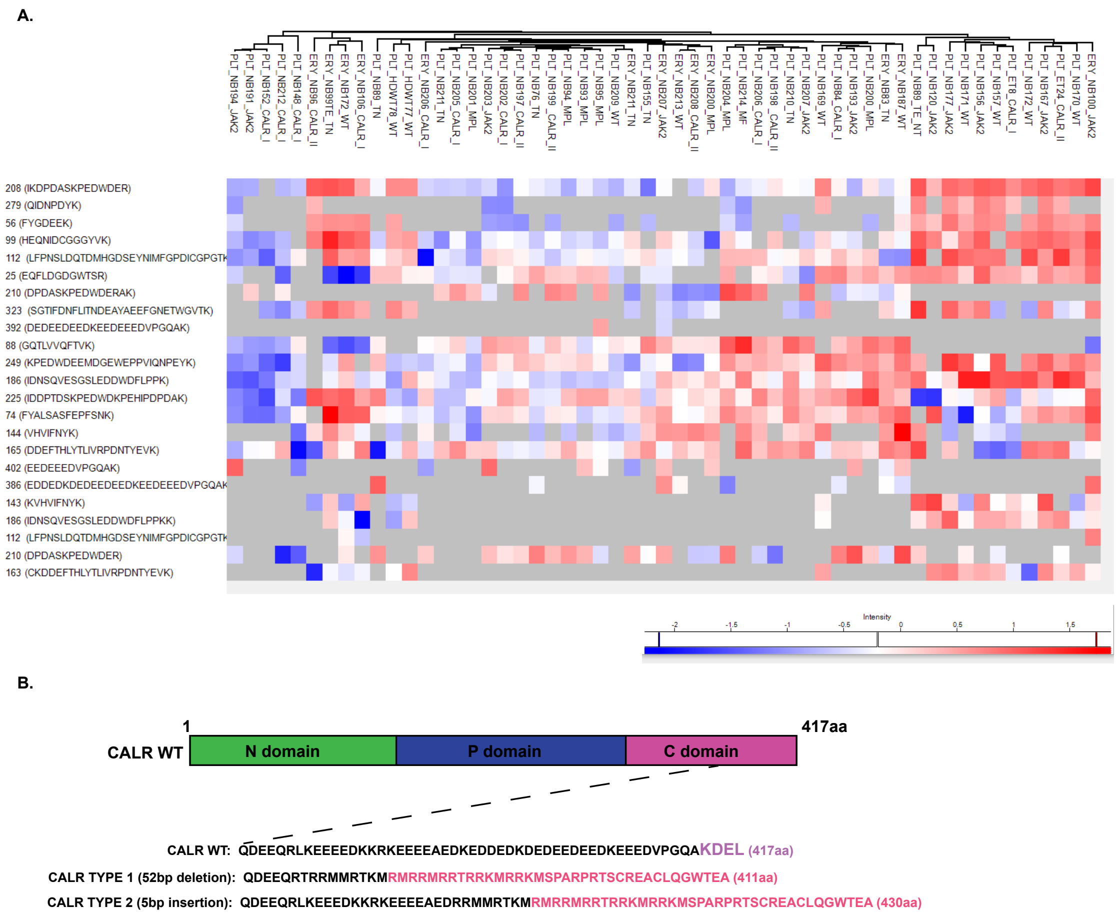


### Supplementary Figure 2

**A.** CALR peptides as detected by LC-MS/MS (1st round of MS) in platelets & erythroid cells from ET patients and normal healthy controls (WT). The sequence of the peptides and the position of the first amino acid in the protein is shown on the left. Grey background indicates lack of detection of the peptide, while dark blue to dark red indicates low to high protein abundance. Specific peptides including the CALRwt (endogenous) are less often detected (e.g. EDDEDKDEDEEDEEDKEEDEEEDVPGQAK) or not detected at all as it is the case for the CALRmut specific peptides (pink colored amino acid sequence is the novel common C-terminal domain in both mutant isoforms, the 52bp deletion and the 5 bp insertion**(B)**). Similar results were obtained from two independent label free mass spectrometry experiments.

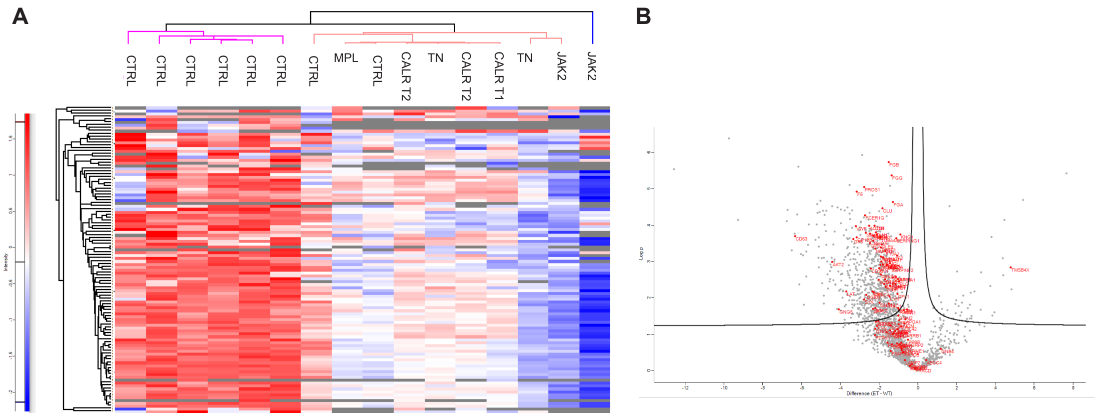


**Supplementary Figure 3**

**A.** Hierarchical clustering of mass spectrometry processed data (Perseus software) from platelet lysates of healthy controls (CTRL) and ET patients. Filtered analysis for platelet activation and aggregation set of proteins (reference protein group) performed on untreated (naïve) platelet samples (CTRL and ET) only. Intensity of the color indicates protein abundance from lowest (dark blue) to highest levels (dark red) for each protein (represented in every line of the heatmap). The majority of the proteins from the reference group had lower abundance in the ET samples **B**. Volcano plot on the same group of proteins showing the significant differences in protein abundance between CTRL and ET platelets after Benjamini-Hochberg t-test (FDR ≤0.05).

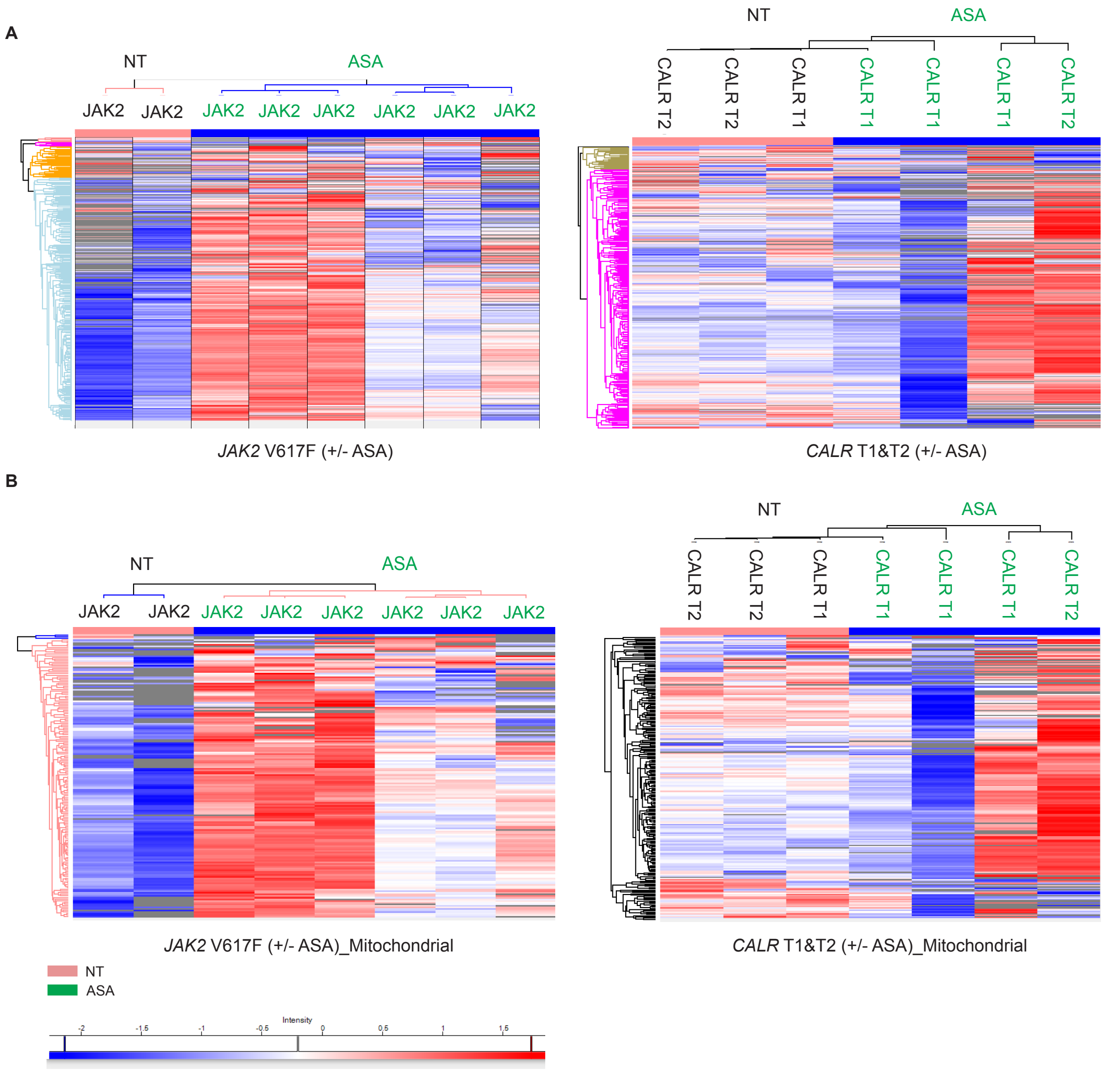


**Supplementary Figure 4**

**A.** Hierarchical clustering of MS processed samples and analyzed by the Perseus Software (See Methods, section Mass Spectrometry) on the effect of acetylsalicylic acid (ASA) on *JAK2* V617F and CALR mutant platelets **B**. Same analysis as in (**A)** but filtered for mitochondrial proteins only. A similar clustering is observed for the *JAK2* V617F samples (+/- ASA) in (**A**) and (**B**) with increased abundance in most of the mitochondrial proteins in all ASA-treated samples. A higher degree of variability is encountered in *CALR* Type1&Type2 platelet samples. Still, ASA-treated *CALR* Type1 (2 out of 4 samples) showed a rather moderate reduction in almost all mitochondrial proteins. However, one *CALR* Type1 and one *CALR* Type2 sample showed an increase in protein abundance in bulk and mitochondrial proteins upon ASA treatment (NT: Non-treated, ASA: acetylsalicylic acid).

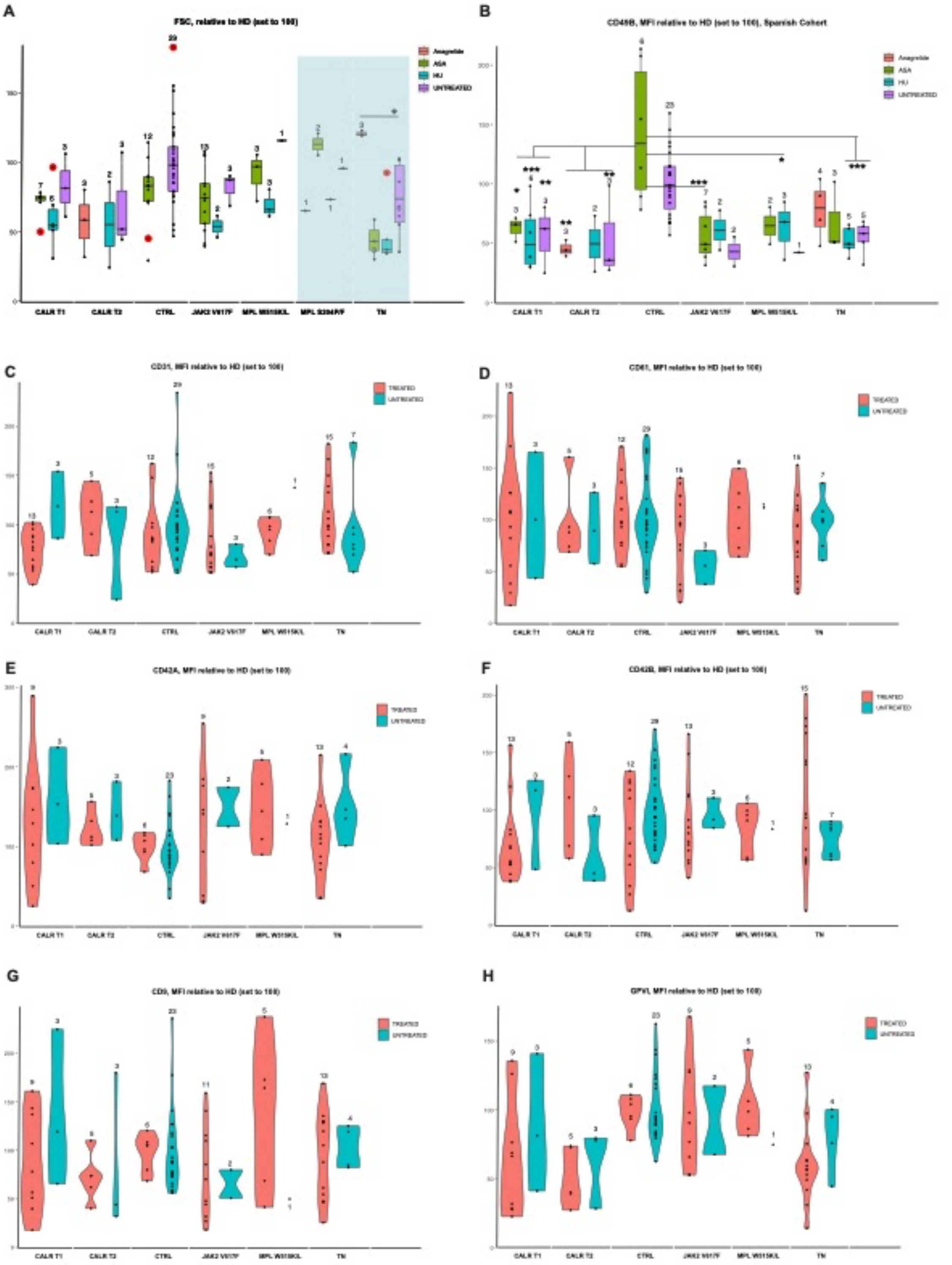


**Supplementary Figure 5**.

**A.** Relative expression of surface markers (Mean fluorescence intensity, MFI) of forward scatter (FSC) and (**B-H**) CD49B, CD31, CD61, CD42A, CD42B, CD9, GPVI of CTRL and ET platelets as calculated by flow cytometry. Untreated and treated conditions (ASA, HU, ANG, or a combination of them) have been plotted except from FSC (**A**) and

CD49B (**B**) that each treatment has been plotted separately per mutational group and CTRL. Treated samples (red violin plots) include all patient treatments (i.e. ASA, ANG, HU, or a combination of them) except from CTRL and *JAK2* V617F groups in which all treated samples are only ASA-treated. The number of analyzed samples per group is indicated on top of each violin plot. Differences are shown when statistical significance was reached. Equal or more than 3 samples per group were analyzed (*p-value ≤0.05, **p-value ≤0.01, ***p-value ≤0.001), ASA: acetylsalicylic acid, HU: hydroxycarbamide, ANG: anagrelide).

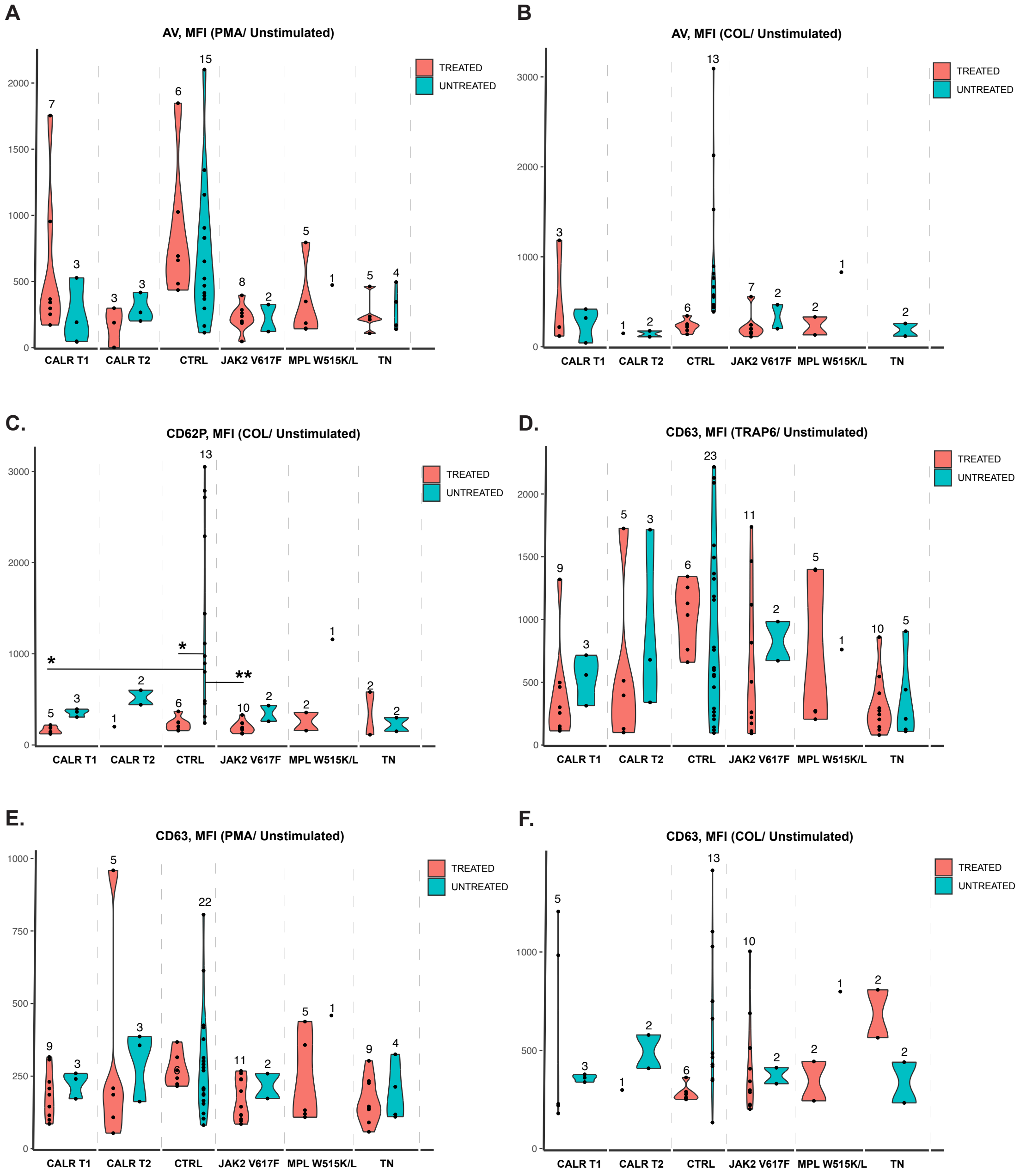


**Supplementary Figure 6**

Ratio of relative expression of surface markers (Mean fluorescence intensity, MFI) obtained by flow cytometry upon agonist stimulation and the unstimulated condition **A.** Ratio of Annexin V (AV), an apoptotic marker upon PMA and **B.** Upon collagen stimulation **C.** CD62P expression levels (ratio) upon collagen stimulation and CD63 expression levels upon TRAP6, PMA, and collagen stimulation **(D-F)**. Specifically, collagen stimulation produced more cell death (debris) at 5min when expression levels

(MFI) of AV, CD62P and CD63 were measured and therefore the number of included samples per mutational group was lower than usual. Treated samples (red violin plots) include all patient treatments (i.e. ASA, ANG, HU, or a combination of them) except from CTRL and *JAK2* V617F groups in which all treated samples are ASA-treated. The number of samples per group is indicated on top of each violin plot. Statistical differences are shown only when significant and only when at least 3 samples per group were analyzed (*p-value ≤0.05, **p-value ≤0.01, ***p-value ≤0.001, ASA: acetylsalicylic acid, HU: hydroxycarbamide, ANG: anagrelide).

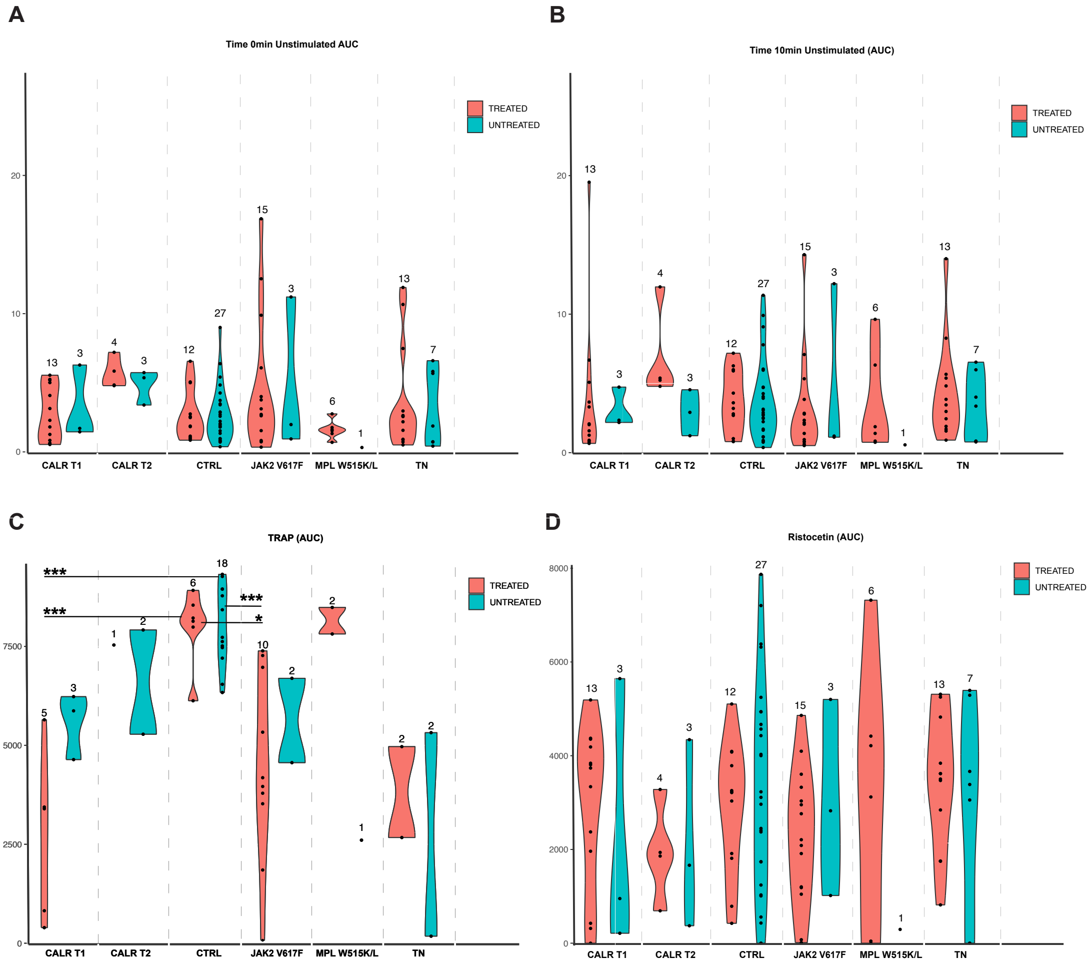
A
Time 0min Unstimulated AUC
TREATED
UNTREATED
CALR T1
CALR T2
CTRL
JAK2 V617F
MPL W515K/L
TN
B
Time 10min Unstimulated (AUC)
C
TRAP (AUC)
***
***
***
*
D
Ristocetin (AUC)

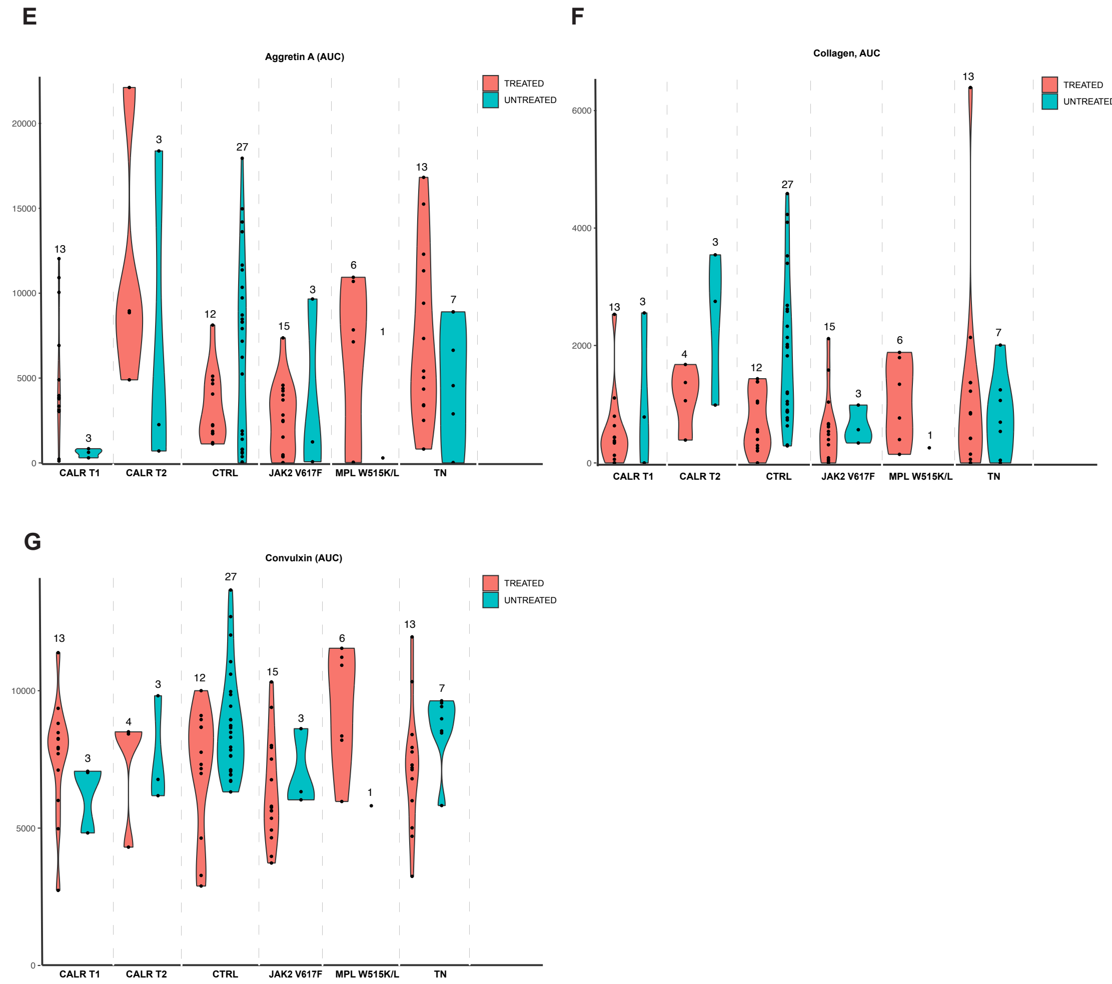


**Supplementary Figure 7**

Area under the curve (AUC) calculated from the time course stimulation of platelets (ET and CTRL) **A.** Time 0min **B.** Time 10min before (**A&B**) and after stimulation (10min max) with different agonists indicated on top of each plot (**C-G**). Treated samples (red violin plots) include all patient treatments (i.e. ASA, ANG, HU, or a combination of them) except from CTRL and *JAK2* V617F groups in which all treated samples are ASA-treated. The number of analyzed samples per group is indicated on top of each violin plot. Statistical differences are shown only when significant and only when at least 3 samples per group were analyzed (* p-value ≤0.05, ** p-value ≤0.01, *** p-value ≤0.001, ASA: acetylsalicylic acid, HU: hydroxycarbamide, ANG: anagrelide).

### Methods (Antibodies)

| Antibody | Cat number | Source | Host | Assay |
|---|---|---|---|---|
| CS | CS14309s | CELL SIGNALING | Rabbit | Western blotting |
| ACO2 | ab110321 | ABCAM | Mouse | Western blotting |
| OGDH | HPA020343 | SIGMA | Rabbit | Western blotting |
| SDHB | ab14714 | ABCAM | Mouse | Western blotting |
| COXIV | CS11967 | CELL SIGNALING | Mouse | Western blotting |
| PKM2 | CS4053S | CELL SIGNALING | Rabbit | Western blotting |
| ACLY | HPA022434 | SIGMA | Rabbit | Western blotting |
| GAPDH | CS2118 | CELL SIGNALING | Rabbit | Western blotting |
| SDHA | ab14715 | ABCAM | Mouse | Western blotting |
| TIM23 | 611223 | BD | Mouse | Western blotting |
| HK2 | CS2867 | CELL SIGNALING | Rabbit | Western blotting |
| PFKP | CS12746 | CELL SIGNALING | Rabbit | Western blotting |
| CD36 | CS14317 | CELL SIGNALING | Rabbit | Western blotting |
| p38 | CS8690 | CELL SIGNALING | Rabbit | Western blotting |
| p-p38 | CS4511 | CELL SIGNALING | Rabbit | Western blotting |
| Tubulin | CS2148 | CELL SIGNALING | Rabbit | Western blotting |
| CD31-APC | 303112 | BIOLEGEND | Mouse | FLOW CYTOMETRY |
| CD31-PE | 303106 | BIOLEGEND | Mouse | FLOW CYTOMETRY |
| CD36-APC | 550956 | BD | Mouse | FLOW CYTOMETRY |
| CD49B-PE | 555669 | BD | Mouse | FLOW CYTOMETRY |
| CD62P-PE | 555524 | BD | Mouse | FLOW CYTOMETRY |
| CD41-FITC | 555748 | BD | Mouse | FLOW CYTOMETRY |
| CD42A-PE | 558819 | BD | Mouse | FLOW CYTOMETRY |
| CD42B- APC | 551061 | BD | Mouse | FLOW CYTOMETRY |
| CD9-FITC | 555371 | BD | Mouse | FLOW CYTOMETRY |
| GPVI-PE | 565241 | BD | Mouse | FLOW CYTOMETRY |
| CD63-APCCY7 | 561982 | BD | Mouse | FLOW CYTOMETRY |
| CD61-FITC | 555753 | BD | Mouse | FLOW CYTOMETRY |
| PAC1-FITC | 340507 | BD | Mouse | FLOW CYTOMETRY |
| 7AAD | 559925 | BD | | FLOW CYTOMETRY |
| ANNEXIN V-V45( | 560506 | BD | | FLOW CYTOMETRY |

Different conjugates have been used on flow cytometry antibodies of the same clone.
